# A case report on gendered biases in a Finnish healthcare AI assistant

**DOI:** 10.64898/2026.04.09.26350383

**Authors:** Rami Luisto, Karoliina Snell, Ville Vartiainen, Enni Sanmark, Sami Äyrämö

## Abstract

In this study, we investigate gender bias in a Retrieval-Augmented Generation (RAG) based AI assistant developed for Finnish wellbeing services counties. We tested the system using 36 clinically relevant queries, each rendered in three gendered variants (male, female, gender-neutral), and evaluated responses using both an LLM-as-a-judge approach and a human expert panel consisting of a physician and a sociologist specializing in ethics.

We observed substantial and clinically significant differences across gendered variants, including differential treatment urgency, inappropriate symptom associations, and misidentification of clinical context. Female variants disproportionately framed responses around childcare and reproductive health regardless of clinical relevance, reflecting societal stereotypes rather than medical reasoning. Bias manifested both at the LLM generation stage and the RAG retrieval stage, in several cases causing the model to hallucinate responses entirely. Some bias patterns were persistent across repeated runs, while others appeared inconsistently, highlighting the challenge of distinguishing systematic bias from stochastic variation.

## 1 Introduction

The use of AI in the domain of healthcare is increasing rapidly [34, 33]. The deployment of AI-powered systems in healthcare raises questions about how these systems treat different patient populations [11, 6, 25]. Any sort of bias in clinical decision support, when present, could lead to differential quality of care with serious consequences for patient outcomes.

One of the main issues when working with modern language models, especially Large Language Models (LLMs), is the “stochastic parrot” effect [2]. Language models tend to regurgitate text and ideas seen in their training data. Since their training data typically consists in large parts of considerable subsets of the whole internet, this tends to also include all kinds of biases. Indeed, Marinucci et al in [21] discuss how various biases, harmful and otherwise, are a natural part of human thought and thus present in any communication. Biases do not arise solely from individual thought processes, but are also shaped and reinforced by broader social structures, historical contexts, and power relations within society, [18]. The biases are then naturally reflected in modern models. Healthcare data in particular has been shown to contain several forms of bias, [24, 27]. These biases often arise from unrepresentative datasets in medical research, societal stereotypes about gendered symptoms, or the attitudes of medical professionals. The effect is not limited to simple biases as LLMs are quite prone to learning more complex logical fallacies as well. [26]

Modern language models process text through the manipulation of so called embedding vectors in their latent space, see e.g. [28]. And thus many AI biases can be observed geometrically; Bolukbasi et al have shown [3] that a “male–female” direction can be observed in word embedding vectors, and that this direction correlates in biased ways with other directions like “programmer–homemaker” or “doctor–nurse”. These kinds of geometric quantitative observations can be used to both detect and debias AI systems [3]. While gender bias is by far not the only bias (or ethical issue) with AI systems (see e.g. [8, 25].), it is typically much easier to probe for a simplistic binary “male–female” differences than for more complex representations of gender as non binary or intersectional issues related to e.g. race or socio-economical status. It is, therefore, a useful starting point for probing the inherent biases of the system.

In this work, we demonstrate how an LLM-based healthcare assistant produced often gendered and different medical advice depending on whether the patient was identified as male, female, or gender-neutral. Our primary contribution is demonstrating that such gendered text and biases can easily emerge even with modern LLMs, making active mitigation an essential component of responsible AI deployment in healthcare.

### A note on terminology

Throughout this document, we use gender to refer to the social category indicated by terms such as “man” and “woman” in the query variants. When discussing biological factors that might justify differential medical treatment, we use “sex”.

## 2 Methods

### 2.1 System setup

The system under review was a development version of an AI assistant built for the Wellbeing Services County of Central Finland (HYVAKS) and the South Savo Wellbeing Services County (ELOISA). The projects were coordinated by the Finnish national AI ecosystem for social and health services in 2025. The system was called *the AI-assistant* and was aimed to be a tool for e.g. nurses. The system would, based on a user’s query, retrieve various healthcare documentation and provide guidance based on it. The system we analyze was created during a project that was subsequently aborted for reasons unrelated to the topic of this report. In particular, the AI system was still in the development phase, and the developers had not yet begun to address any possible biases in the system. Thus, we are hesitant to claim that the issues we detect and discuss here would have been present in the final system. Regardless, we believe there is value in reporting these interim findings since they highlight how easily biases can occur in such projects. Especially since this project was made public, and the approach could be replicated by other groups.

The system was built in the Microsoft Azure cloud environment and used the Retrieval Augmented Generation (RAG) architecture [20, 9, 1]. In RAG the system retrieves the most relevant pieces of information related to the user query from given data sources using *vector search*. An embedding model is used to transform the text into a numerical vector whose geometry encodes the semantic content of the text and to create a Vector Database that consists of the embedding vectors, and thus semantic meaning, of all the source documents. An incoming query will then also be mapped to an embedding vector, and the query embedding vector is compared against all the existing vectors in the vector database. The retrieval method used in this system was Microsoft’s Azure Cognitive Search [23]^1^. The retrieved texts are then combined with the system prompt and original user query and passed on to the LLM responsible for creating the final answer. This inclusion of more context then enables the LLM to provide more accurate responses based on specific documents it can also cite.

We emphasize that the system has no persistent memory, i.e. the inputs or results of one query do not effect any other queries.

### 2.2 Evaluations

The 40 test queries produced during the first round human testing were assessed for amenablity to gendering. For example, “miesten virstaongelmiin jumppaohjeita” (exercise instructions for men’s urinary problems) was deemed unsuitable, as urological issues can differ considerably between sexes, whereas “Korvan tukkoisuus” (ear congestion) should be more gender-stable. From the original 40 queries, 36 were deemed suitable for gendering.

We then generated gendered variants for these. As an example, below are the three variants generated from the query “Korvan tukkoisuus” (ear congestion)^2^.

- Miehen korvan tukkoisuus. (“Ear congestion in a man.”)
- Naisen korvan tukkoisuus. (“Ear congestion in a woman.”)
- Henkilön korvan tukkoisuus. (“Ear congestion in a person.”)

We ran these triplets of gendered variants through the system three times with a few variations to the procedure.

1. As a baseline we simply ran the system with the same configuration as the production system. This is our “main variation”.
2. To understand the role of the particular language model used in the system, gpt-4o-mini, we then ran the same study but using the more modern gpt-5-mini model as the LLM.
3. Finally we ran a version to test the effect of the RAG system’s retrieval stage to the results by first running the queries against Azure Cognitive Search and then using the gpt-4o-mini model to formulate the response, but instead of using as context the documents corresponding to the particular gendered variants, we would use either of:
  a. The documents found with the *neutral* variant.
  b. The combined set of documents found by *any* of the three gendered variants.

For these results we first assessed the biases by using an LLM-as-ajudge approach where another LLM (OpenAI’s GPT-5) was given the task to assess bias levels on a scale of None, Low, Medium, High. This judgement was ran against the results again in triplicate. For all of the variations 1, 2, 3a and 3b we did a superficial check of the judgements and verified that they seem reasonable on average.

For the “main variation” we also had a medical doctor and a sociologist specializing in ethics go through the results and the LLM-as-a-judge analysis in detail and discuss if any bias or other interesting medical phenomenon were seen. The core idea here was to combine the bias expertise of a sosiologist with the medical expertise of a doctor to better understand which differences were medically significant and reflected existing biases in society. The core questions they asked were as follows:

- Are the differences between the gendered variants’ answers gendered or simply random variations?
- Are there medically valid reasons for the observed differences?
- Is there a difference between the bias level judgements of the LLM-as-a-judge and the human panel?

## 3 Results

We observed substantial variance across queries. For some queries, there was little difference between the gendered versions, while others showed clear changes in medical advice or response urgency. The presence of bias was not always persistent; as mentioned earlier, we ran the queries three times and some biases persisted across all runs while other differences appeared inconsistently. The bias patterns were moderately stable (Table 1).

**Table 1:** Distribution of bias levels across three runs.

| Bias Level | Run 1 | Run 2 | Run 3 |
| --- | --- | --- | --- |
| None | 8 | 12 | 9 |
| Low | 4 | 2 | 4 |
| Medium | 15 | 15 | 15 |
| High | 9 | 7 | 8 |

When examining the bias evaluations across the three runs for the same query, the most common combinations are presented in Table 2.

**Table 2:** Bias consistency patterns across three runs (N=36 queries)

| Pattern across runs | N | % | Interpretation |
| --- | --- | --- | --- |
| 1× medium, 2× none | 8 | 22 | Low bias |
| 2× medium, 1× none | 4 | 11 | Low bias |
| 3× high | 4 | 11 | High bias |
| 3× medium | 4 | 11 | Medium bias |
| 3× none | 2 | 6 | No bias |
| Other patterns | 14 | 39 | Mixed |

The remaining 38.9% of queries (14 queries) fell into various other combinations involving low bias levels or mixed high/medium patterns.

According to our panel of a medical doctor and a sociologist evaluating the bias there was a clear theme where the advice given to women was more often connected to child care. This reflects stereotypical family roles, though there also seemed to be aspects of other stereotypes related to “hormones” or social pressures. There were also results which were gendered for medically valid reasons, e.g. relating pregnancy to women. The panel also observed a possible difference in the language used to address men or women, especially in the level of being commanding or advising, though the effect was not strong enough to be deemed systematic, at least with our sample size. Finally, we note that the panelists occasionally interpreted the results from different points of view. This not so much a difference in opinion, but rather reflects their distinct professional positionalities [10].

### 3.1 Concrete bias examples

There was a systematic tendency to present women as caretakers of children and family while men were excluded from this role even when querying on family services. For example, when querying on contact information for Maternity and Child Health Clinic (“äitiys- ja lastenneuvola” in Finnish) providing services for prevention, screening and monitoring during pregnancy, and early childhood the female variant received recommendation on contacting the clinic via phone or interned based services without providing the exact contact information. The male variant received contact information for services providing social support for families (“Perheneuvola” in Finnish). Additionally, when the system was queried for instructions on patient safety incidence report the female variant was given instructions for child protection services. The theme continued also when seeking information on medical procedures. When querying on “the snowman test”, which is a collection of tasks to assess the linguistic development of children, the neutral and female variants received mostly correct information on the test while the male variant produced answers such as “the test is not applicable to adults”, gave instructions for a semen test or hallucinated a test procedure where a (male) child is observed in a cold snowy environment.

Finally, one of the most egregious cases involved ear congestion. In the three gendered variants, the male variant includes advise to seek immediate medical care if there is sudden loss of hearing. For the female variant, part of the suggestion is that care should be sought only if the problem persists or is accompanied by lower abdominal pain. The neutral version mostly suggests waiting at home unless issues persist or worsen. This differential is clinically significant. While the gender neutral variant is likely to be the most reasonable advice in most cases, sudden deafness, often caused by sensorineural hearing, loss is a medical emergency with treatment window of 24–72 hours for optimal outcomes with corticosteroid therapy [5]. The fact that this warning appears only in the male variant and is absent from both the female and neutral variants represents exactly the type of differential treatment that could have serious clinical consequences in a deployed system.

The mention of “lower abdominal pain” in the female variant is separately noteworthy. There is no anatomical or physiological reason why ear congestion in women would be associated with abdominal pain [7]. One interpretation is that this pattern reflects documented tendencies in medical practice to attribute women’s symptoms to reproductive or gynecological causes, [12], which again relates to the theme of associating women with reproduction. In this particular case, the male response was also notably longer than the others, but statistically, the responses were the same length across all 36 queries (see Table 3).

**Table 3:** Response length statistics (word count) across gendered variants for all 36 queries.

| Metric | Male | Female | Neutral |
| --- | --- | --- | --- |
| Mean | 259.6 | 262.2 | 266.5 |
| Median | 273.0 | 284.0 | 271.5 |
| Std Dev | 77.1 | 80.2 | 78.1 |
| Min | 94 | 76 | 64 |
| Max | 378 | 395 | 395 |

Most of the queries such as “aivojen pet-tt tutkimukseen valmistautuminen” (brain PET-CT preparation) and “äkillinen kova selkäkipu, iskias” (sudden severe back pain, sciatica) resulted in similar medical checklists across the variants with only minor stylistic differences.

While the clearly biased events were more of an exception than a rule, easily finding gendered differences in just 36 examples does point towards a major issue, especially when combined with the volatility of responses between the runs. Though a part of the challenge in this analysis was that even when no bias was present, the answers might vary quite a lot between identical queries. Detecting consistent bias from random noise effects then turns challenging.

### 3.2 Retrieval and LLM stage variations

When running variants of our queries where we altered the set of documents that the LLM model was given for context, the distribution of bias levels as measured by the LLM-as-a-judge was almost identical to the distribution observed in the standard variation. Despite the lack of significant differences in the levels of detected bias, there was one clear outlier. The query “Mikä on VOITAS?” (What is VOITAS?) was particularly susceptible to gendered versions in the data retrieval stage. Here VOITAS is a fall-prevention program designed for the elderly. When the model answered this query based on the neutral RAG document retrieval, the results were as expected, but the gendered versions essentially hallucinated answers; for men it discussed about a non-existing men’s wellbeing program, while for women it discussed about substance abuse during pregnancy. (Note again the connection between women and pregnancy/children.) The underlying reason here seems to be that the male and female variants produced no VOITAS related documents in the retrieval stage, with the gender triggers dominating what documents were retrieved. Without relevant material returned from the RAG stage, the model opted to make something up i.e. hallucinate. For comparison, a purely administrative query for the Kyllö healthcare center contact information also produced biased results where the female variant was not given specific contact information even though data retrieved was largely stable between variants. In particular, the correct contact information was among the retrieved data for each gendered variant, but the male variant of the query was more consistently provided with the proper contact information.

We also ran an analysis where we changed the gpt-4o-mini LLM from the evaluation stage into a more modern gpt-5-mini. The levels of bias were slightly reduced according to the LLM-as-a-judge but clear bias still remained.

## 4 Discussion

### 4.1 Possible causes

We identify several potential causes for the observed bias patterns, though we do not claim this list to be exhaustive. Understanding possible causes such as these is important for designing appropriate mitigation strategies.

#### LLM inherent bias

Our primary hypothesis is that the underlying language model carries gender biases from its training data. Large language models trained on internet-scale corpora inevitably absorb societal biases present in those texts [2, 21]. These biases can manifest as differential treatment recommendations or urgency assessments when gender markers are present in the input.

#### Vector search bias

The embedding model used for retrieval may encode gender stereotypes in its vector representations. Previous research has demonstrated that word embeddings can associate certain conditions or treatments more strongly with one gender [3]. However, our results do not seem to support this; at least the effect here is less prominent compared to the biases produced at the generation phase. Regardless, with our qualitative approach we are not ready to make strong statements on the strength of this effect. See also [13] for further discussion on this topic.

#### Term dominance in short queries

Many of our test queries were relatively brief and in short queries, individual words carry more semantic weight. When we add a gendered term like “miehen” (man’s) or “naisen” (woman’s) to a two-word query, that gendered term may disproportionately influence both the embedding vector and the retrieved documents. Though we note that the short query was always combined with a larger base prompt. Regardless, longer test texts would be fruitful here, see also again [31, 36].

### 4.2 Possible solutions

There is considerable literature on the best practices of bias mitigation; see e.g. [30, 17, 22, 31, 36] and references within. Note in particular that [30] has also evaluation source code available, and [17] talks specifically about bias in medical RAG pipelines. While a thorough survey of evaluation and mitigation strategies is beyond the scope of this observational study, we list a few practical approaches that practitioners might consider. This listing of concrete mitigation strategies is partially motivated by Isaksson [15] who notes the tendency of academic papers to lack in giving concrete actions for Healthcare AI bias mitigation. In this spirit we also strongly recommend that the topic of (gender) bias, both its detection and mitigation, is taken up in any project design, and a concrete responsibility is given e.g. to the project manager or steering committee to make sure that this is tested throughout development.

We also warrant deep caution with these kinds of systems. With various types of bias tests we can easily detect when clear bias is present, but negative test results do not prove that bias is *not* present in some form. Furthermore, as is argued in [21], various biases seem to be part of the human thought process and thus there it might be impossible to completely remove biases from a system that can approximate human linguistical skill to any reasonable degree.

All that being said, for more technical approaches to bias mitigation we suggest the following.

#### Query preprocessing

One approach involves pre-processing queries before they enter the RAG pipeline. This could involve extending short queries with additional context to dilute the influence of gender/sex markers, or systematically removing any such information which is not medically relevant to the query. The challenge here is naturally to identify which cases are such that e.g. gender/sex information can be omitted. For example, the risk for cardiovascular disease is markedly different between sexes, [32].

#### Multi-variant querying

A more robust approach might involve running multiple query variants (male, female, neutral) through the system and aggregating or comparing results. Significant divergence between variants could trigger human review or a more conservative response. A challenge here is that this approach can grow infeasible if there are many different “bias targets” like sex/gender, race, age, etc. that we need to debias against. For example [16] comments on the exponential growth of intersectional subgroups one might have to evaluate.

#### Post-processing bias detection

A secondary model could analyze generated responses for potential biases before presenting them to users. Such a system could work particularly well when combined with a system that runs different variants through the LLM. A major question then is on whether we can trust this post-processing LLM to be unbiased? See [35] for a less flattering study on LLMs as judges in the healthcare sector, and [14, 29] for a propositions on how to implement such approaches.

#### Use (medical) professionals throughout the development

Biases manifest differently depending on one’s perspective, which makes their detection nontrivial. This can be mitigated with epistemological diversity, i.e. by utilizing diverse epistemic positions as much as possible during the full development cycle of these kinds of systems. This should include not only medical professionals, but also patients, ethicists, guideline creators, and service system experts [19, 4].

## 5 Conclusion

We documented systematic and persistent differences in medical advice across gendered query variants in a RAG based healthcare assistant. In one notable example involving ear congestion, the male variant received time-sensitive warnings about sudden hearing loss, a medical emergency with a 24–72 hour treatment window, while the female variant received advice that inappropriately linked the condition to abdominal pain. Our observations demonstrate that gender biases arise quite easily in RAG-based systems without active mitigation efforts. We furthermore suspect that such direct gender biases are only the tip of an iceberg. More nuanced issues, such as intersectional biases involving several demographic factors or biases in treatment intensity recommendations, can be much harder to probe, analyze, or mitigate. We suggest that developers of healthcare AI systems should incorporate bias testing as a standard component of development, not an afterthought. We also emphasize here that with the current reliance on models where we do not have full visibility to the model contents or training data, the root causes for biases are hard to address. We can build add-on systems to detect and mitigate the issue, but for a fully unbiased system we would need a whole new paradigm for data gathering and model training, which seems to be out of the scope of possibility for the field at the moment.

## Data Availability

All data produced in the present study are available upon reasonable request to the authors

## Conflicts of Interest

During the majority of this work the first author was employed at Digital Workforce Services (DWF), but the totality of this work was carried out independently of the company as a researcher at the University of Jyväskylä and the Helsinki University Hospital. We also note that DWF, despite providing services in this specific field, was not involved in the development of the AI assistant project discussed here. All the parties involved in the project and its analysis were made aware of the first author’s employment at DWF before the start of the project.

## Footnotes

1 We note that the Azure Cognitive Search builds upon the simple vector search approach with further techniques which are not germane to the discussion here.

2 We note that these variants are linguistically more fluent in Finnish than in English. Possibly partially due to the fact that Finnish has no gendered pronouns, and thus when gender needs to be specified it relies on direct identifiers like “man” or “woman”.

## Notes

### Competing Interest Statement

RL was employed Digital Workforce Plc when working on this project and owns the private research company IatroData Ltd. VV has received unrelated consultation fees and honoraria from Orion Pharma (Espoo, Finland).

### Funding Statement

This study did not receive any funding.

